# Lymphoid Enhancer-Binding Factor 1 (LEF1) immunostaining as a surrogate of β-catenin (*CTNNB1)* mutations

**DOI:** 10.1101/2022.03.30.22273113

**Authors:** Ekkehard Hewer, Pascal Fischer, Erik Vassella, Laura Knabben, Sara Imboden, Michael D. Mueller, Tilman T. Rau, Matthias S. Dettmer

**Affiliations:** Institute of Pathology, University of Bern, Bern, Switzerland; Institute of Pathology, Lausanne University Hospital and University of Lausanne, Lausanne, Switzerland; Institute of Pathology, Heinrich-Heine-University and University Hospital Duesseldorf, Düsseldorf, Germany; Department of Gynaecology and Obstetrics, Inselspital University Hospital, Bern, Switzerland; Institute of Pathology, Klinikum Stuttgart, Stuttgart, Germany

**Keywords:** β-catenin, lymphoid enhancer-binding factor 1, WNT signaling

## Abstract

**Background:** Activating mutations affecting exon 3 of the β-catenin (*CTNNB1*) gene result in constitutive activation of WNT signaling and are a diagnostic hallmark of several tumor entities including desmoid-type fibromatosis. They also define clinically relevant tumor subtypes within certain entities such as endometrioid carcinoma. In diagnostics, β-catenin immunohistochemistry is widely used as a surrogate for *CTNNB1* mutations, but is often difficult to assess in practice, given that the characteristic nuclear translocation may be focal or hard to distinguish from spillover of the normal membranous staining.

**Study design and methods:** We therefore examined Lymphoid Enhancer-Binding Factor 1 (LEF1) immunostaining, a nuclear marker of WNT activation that serves as a potential surrogate of *CTNNB1* mutations.

**Results:** In a cohort of endometrial carcinomas (n=255) LEF1 predicted *CTNNB1* mutations correctly in 85%, while β-catenin was 76% accurate. Across a variety of entities characterized by *CTNNB1* mutations as putative drivers, we found diffuse and strong expression of LEF1 in 77% of cases. LEF1 immunostaining proved easier to interpret than β-catenin immunostaining in 54% of cases, more difficult in 1% of cases, and comparable in the remaining cases.

**Conclusion:** We conclude that LEF1 immunostaining is a useful surrogate marker for *CTNNB1* mutations. It favorably complements β-catenin immunohistochemistry and outperforms the latter as a single marker.

## Introduction

Mutations in exon 3 of the *CTNNB1* gene, which encodes β-catenin, lead to constitutive activation of WNT signaling, contributing to tumorigenesis. They are recognized as the molecular hallmark of various tumor types (including desmoid-type fibromatosis, pilomatricomas, adamantinomatous craniopharyngiomas). Under physiological conditions β-catenin attaches to adhaerens junctions, resulting in a membranous staining, while activating *CTNNB1* mutations result in nuclear translocation of β-catenin protein. This nuclear translocation can be visualized by immunohistochemistry, as *CTNNB1* mutated tumors show combined membranous and nuclear β-catenin staining.

In routine diagnostic practice, β-catenin immunostaining is widely used as a surrogate marker for *CTNNB1* mutations in a variety of lesions because it is less expensive and can be performed more quickly than gene sequencing. Diagnostic utility of β-catenin immunostaining has e.g. be demonstrated in solid pseudopapillary neoplasm of the pancreas, where it can be particularly helpful in the diagnosis of specimens with limited material obtained by endoscopic ultrasound-guided fine needle aspiration [1]. Similarly, in desmoid-type fibromatosis, it has been shown that identification of nuclear β-catenin staining as a surrogate of *CTNNB1* mutations can be used diagnostically in difficult lesions [2,3].

Sporadic endometrial carcinomas harbor *CTNNB1* mutations in about 20% [4]. Recently, these mutations were reported to identify patients at increased risk of recurrence, and in a follow-up work, the same group reported that β-catenin immunostaining can be used as a screening tool to identify tumors that should undergo *CTNNB1* sequencing [4,5]. This was confirmed in a large meta-analysis which found that β-catenin immunostaining can indeed be used as a screening tool for *CTNNB1* mutations [6].

Currently, more than 60’000 new cases of endometrial cancer are diagnosed in the United States each year, according to the SEER database, representing 7% of all new cancer cases diagnosed in women [7]. Incidence and mortality rates are increasing, making it urgent to identify patients at risk of tumor recurrence or progression.

Because of the often focal nature of nuclear translocation and nuclear spillover of membranous staining, β-catenin immunohistochemistry is notoriously difficult to evaluate and requires considerable expertise [8]. Therefore, a new marker that is easier to interpret would be very helpful. This prompted us to investigate the utility of LEF1 (Lymphoid Enhancer Binding Factor 1), a downstream mediator of WNT signaling. The *LEF1* gene belongs to the TCF/LEF (T cell factor/lymphoid enhancer factor) gene family. LEF1 physically interacts with β-catenin when the latter is translocated to the nucleus and mediates WNT signaling. LEF1 is currently used as a diagnostic marker for chronic lymphocytic leukemia [9] and is therefore widely available across diagnostic immunohistochemistry laboratories. Here, we assess its potential as an immunohistochemical surrogate for *CTNNB1* mutations.

## Material and methods

### Case selection and tissue microarray construction

A tissue microarray (TMA) was constructed as previously described [10]. All clinical and histopathological data of the cohort were previously described [11]. Three representative punches (1mm core) were taken per carcinoma to address possible intratumoral heterogeneity, and two additional punches were taken for DNA extraction.

We furthermore identified tumor samples across a variety of tumor types in which sequencing within our department’s diagnostic routine for diagnostic or predictive purposes had shown a pathogenic *CTNNB1*.

The study was conducted in accordance with the Swiss Federal Law on Research Involving Human Subjects and with the approval of the Ethics Committee of the Canton of Bern (KEK 2014-200 and 2017-1189).

### Immunohistochemistry

For cases with endometrial carcinoma of which mutational status was available (n=130; table 1) β-catenin and Lymphoid enhancer-binding factor 1 (LEF1; n=156) immunoexpression were assessed. Immunostainings were performed on a Leica Microsystems Bond Max Stainer. Formalin-fixed, paraffin-embedded slides were stained with β-catenin (CellMarque, clone 14, order nr 224M-15, dilution 1:400) and LEF1 (Abcam, clone EPR2029Y, order nr ab137872, dilution 1:100). Staining was assessed by two experienced pathologists (EH and MSD) who were blinded for clinical data and molecular findings. Any nuclear positivity for β-catenin was considered positive. With regard to LEF1 immunostaining, preliminary analysis of the complete histological slides had shown that the vast majority of tumors were either completely negative or that essentially all tumor cells were strongly stained. Occasional cases (*CTNNB1* wild type) showed focal positivity (usually of heterogeneous intensity) at the invasion front, possibly associated with epithelial-mesenchymal transformation. Therefore, a formal cut-off at 50% tumor cell staining was chosen for LEF1 to be considered positive. For incongruent cases, consensus was reached in a second session. We also assessed the difficulty in deciding whether a stain was positive or negative. The same interpretative criteria were used for LEF1 and β-catenin immunostaining on other tumor types.

**Table 1:** Clinico-pathological characteristics of endometrial carcinoma cases with available *CTNNB1* mutational status.

|  |  |
| --- | --- |
| **Number of Patients** | 130 |
| RFS months (average) ± st dev | 50.1 ± 39.0 |
| DSS (average) ± st dev | 27.8 ± 21.8 |
| OS (average) ± st dev | 55.9 ± 37.3 |
| **FIGO Stage** |  |
| IA | 49 (37.7%) |
| IB | 33 (25.4%) |
| II | 13 (10%) |
| IIIA | 4 (3.1%) |
| IIIB | 3 (2.3%) |
| IIIC1 | 8 (6.2%) |
| IIIC2 | 8 (6.2%) |
| IVB | 12 (9.2%) |
| **Histological tumor grade** |  |
| well differentiated (G1) | 43 (33.1%) |
| moderately differentiated (G2) | 55 (42.3%) |
| poorly differentiated (G3) | 32 (24.6%) |
| **MSI Status** |  |
| stable | 84 (64.6%) |
| aberrant | 46 (35.4%) |
| **POLE Status** |  |
| WT | 116 (89.2%) |
| hotspot | 5 (3.8%) |
| VUS | 9 (6.9%) |
| **TCGA group** |  |
| 1 | 5 (3.8%) |
| 2 | 44 (33.8%) |
| 3 | 57 (43.8%) |
| 4 | 24 n(18.5%) |
| **Mutations in WNT pathway genes** |  |
| *APC* | 1 (0.8%) |
| *AXIN1* | 1 (0.8%) |
| *AXIN2* | 2 (1.5%) |
| *CSNK1A1* | 1 (0.8%) |
| *CTNNB1* | 25 (19.2%) |
| *EP300* | 3 (2.3%) |
| *PPP2R1A* | 7 (5.4%) |
| WT | 90 (69.2%) |

RFS: recurrence-free survival, DSS: disease-specific survival, OS: overall survival, WT: wildtype, VUS: variant of undetermined significance

### DNA extraction

DNA was extracted and subjected to molecular analysis as described elsewhere [12]. Tumor tissue was identified by a molecular pathologist (MSD), and the area of interest was labeled on a slide stained with hematoxylin and eosin (H&E). The area of interest was identified and punched out of the formalin fixed and paraffin embedded tissue (FFPE) block.

### Sanger Sequencing

For sanger sequencing a fragment encompassing exon 3 of the *CTNNB1* gene was amplified by PCR using the primer pair 5’-GCC ATG GAA CCA GAC AG-3’ and 5’-TTC CCA CTC ATA CAG GAC TT-3’ and analyzed by Sanger Sequencing using a Genetic Analyzer (GA3500, Thermofisher).

### Next generation sequencing

64 endometrial cancers were sequenced using Illumina’s TruSight oncology 500 panel (TSO500), which includes 523 genes and allowed us to examine not only *CTNNB1* mutations but also several other genes of the WNT pathway (*APC, AXIN1, AXIN2, CSNK1A1, EP300* and *PPP2R1A*).

### Statistical analyses

Statistical analyses were performed with SPSS (Version 21.0). The Kolmogorov– Smirnov test was used to assess the sample distribution. Statistics were calculated using Chi-square test and Fisher‘s exact test. Cox regression was used for the multivariate analysis and the Kaplan-Meier method for the survival analysis. A P-value of <0.05 was considered to be statistically significant.

## Results

### β-catenin and LEF1 in endometrial carcinomas

*CTNNB1* exon 3 mutations were found in 25 out of 130 cases (19%). Mutational status correlated excellently with immunostaining for β-catenin (p<0.001) as well as for LEF1 (p<0.001). Sensitivity and specificity of β-catenin were 72% and 76%, respectively; for LEF1 they were 64% and 95%, respectively. We found LEF1 easier to interpret in 54% cases (n=70), comparable to β-catenin in 45% of cases (n=58) and more difficult in 1% of cases (n=2) (Fig. 1).

**Figure 1:**
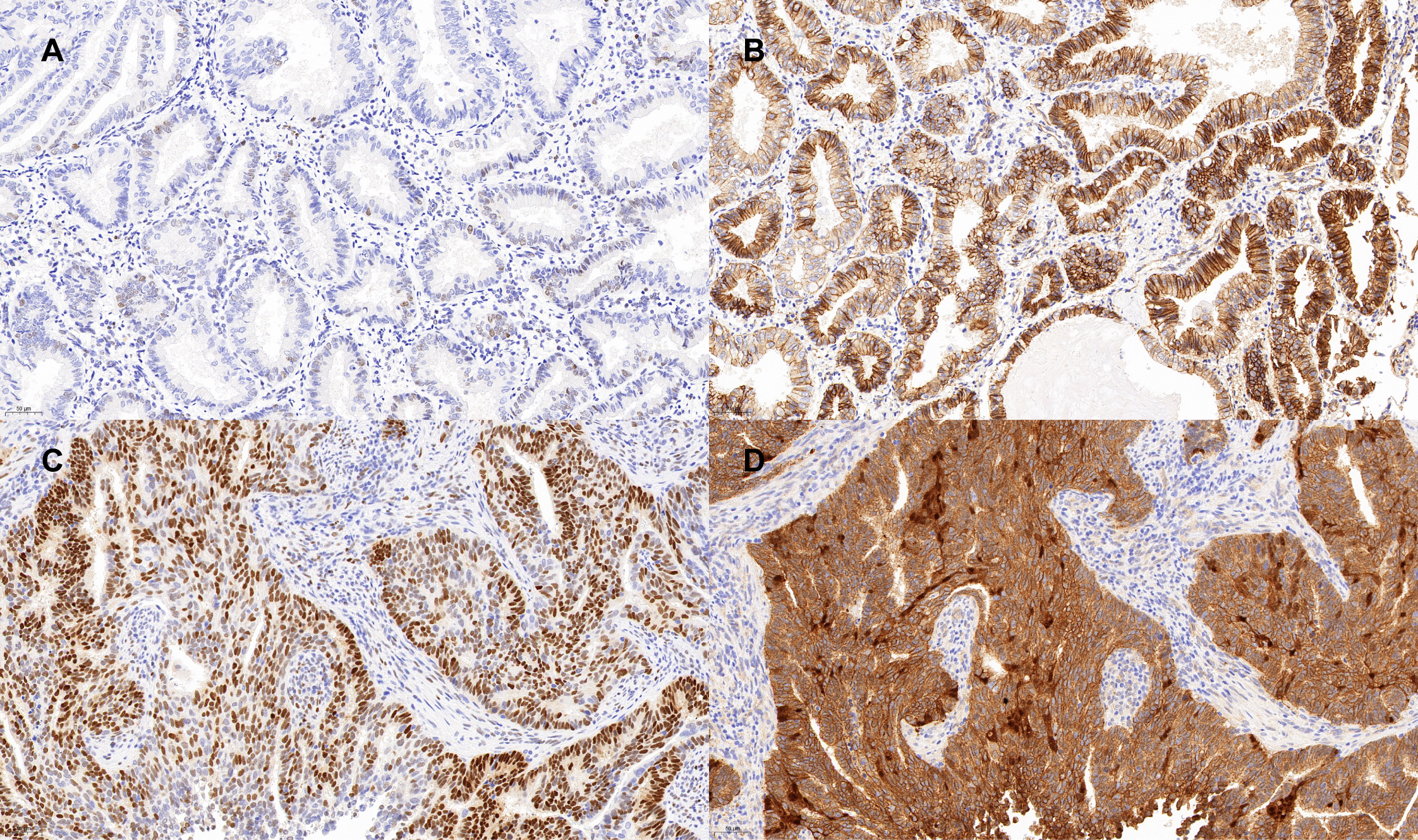
LEF1 and β-catenin in endometrial carcinoma. A: negativity of LEF1; B: membranous positivity of β-catenin and partial nuclear positivity, rendering a classification difficult; C: strong nuclear positivity of LEF1; D: strong nuclear, cytoplasmic and membranous positivity of β-catenin.

β-catenin and LEF1 immunohistochemistry were both predictive of relapse free survival (RFS) (p<0.05) and in addition, LEF1 also predicted overall survival (OS) (p<0.05) in patients with endometrial carcinoma, whereas β-catenin staining did not reach statistical significance (Fig. 2).

**Figure 2:**
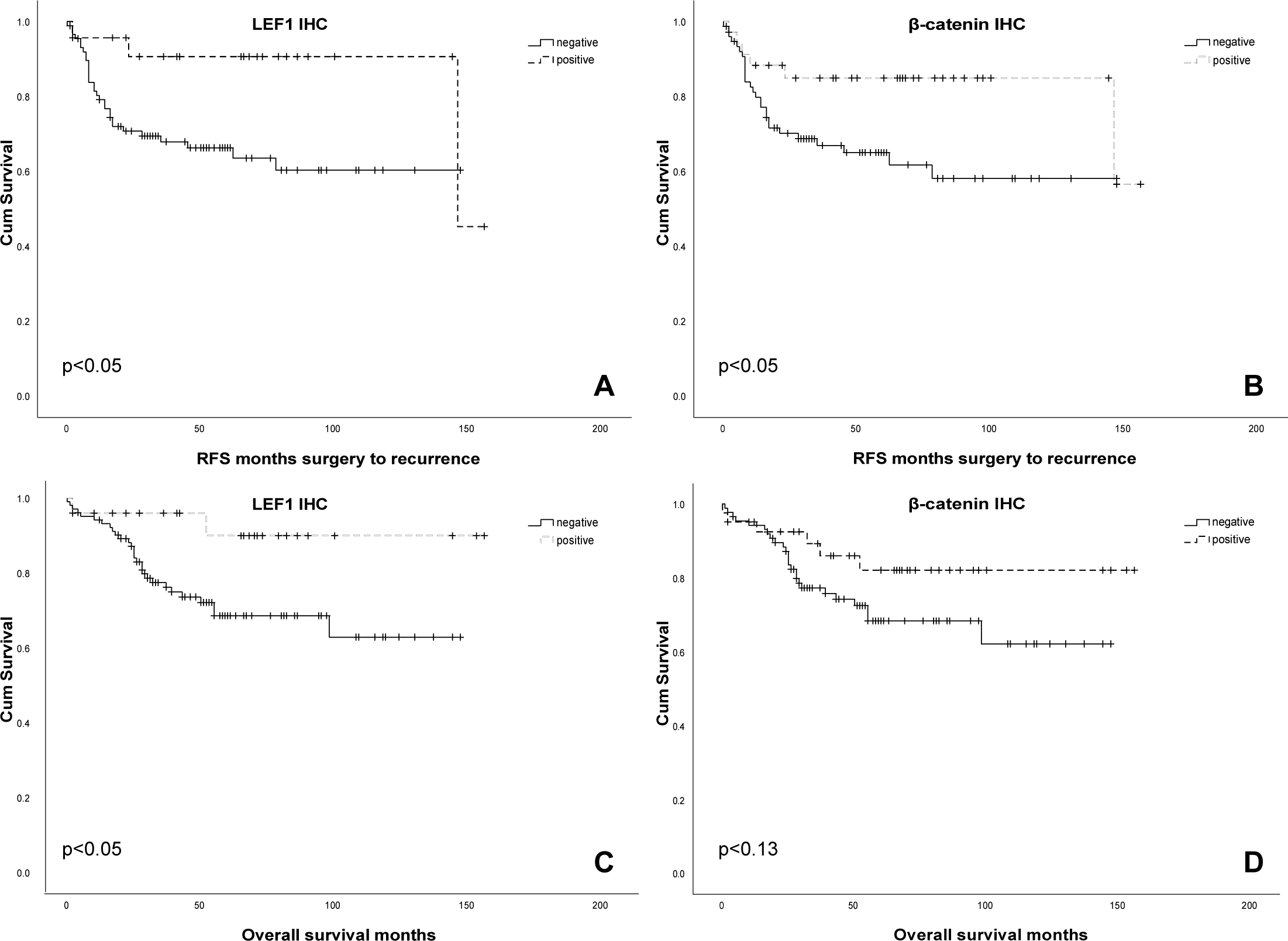
Survival curves LEF1 and β-catenin immunohistochemistry; A: LEF1 relapse free survival (RFS)(p<0.05); B: β-catenin RFS (p<0.05); C: LEF1 overall survival (OS)(p<0.05); D: β-catenin OS (p<0.13).

*CTNNB1* mutations were equally correlated with OS (Fig. 3), a finding which could be confirmed by the TCGA Dataset [13,14] including 542 endometrial carcinomas (Fig. 4). RFS was not predicted by *CTNNB1*, probably due to an outlier (Fig. 3).

**Figure 3:**
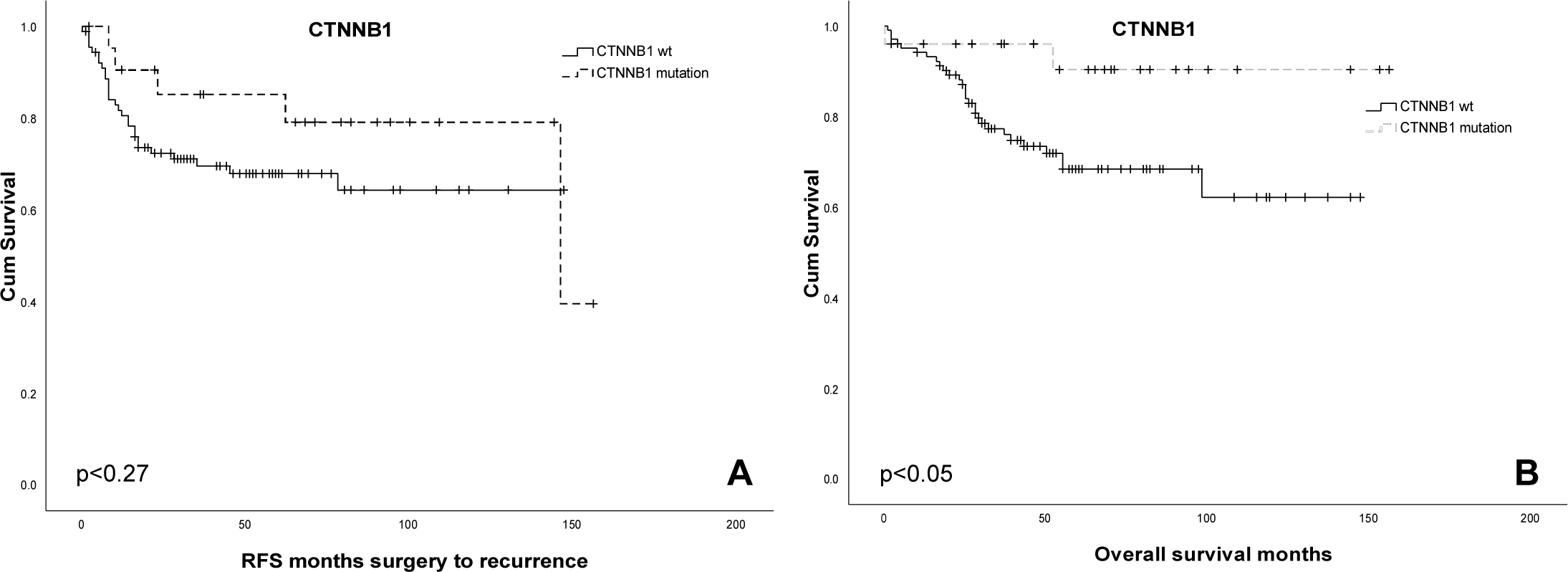
Survival curves *CTNNB1*; A: relapse free survival (p<0.27); B: overall survival (p<0.05).

**Figure 4:**
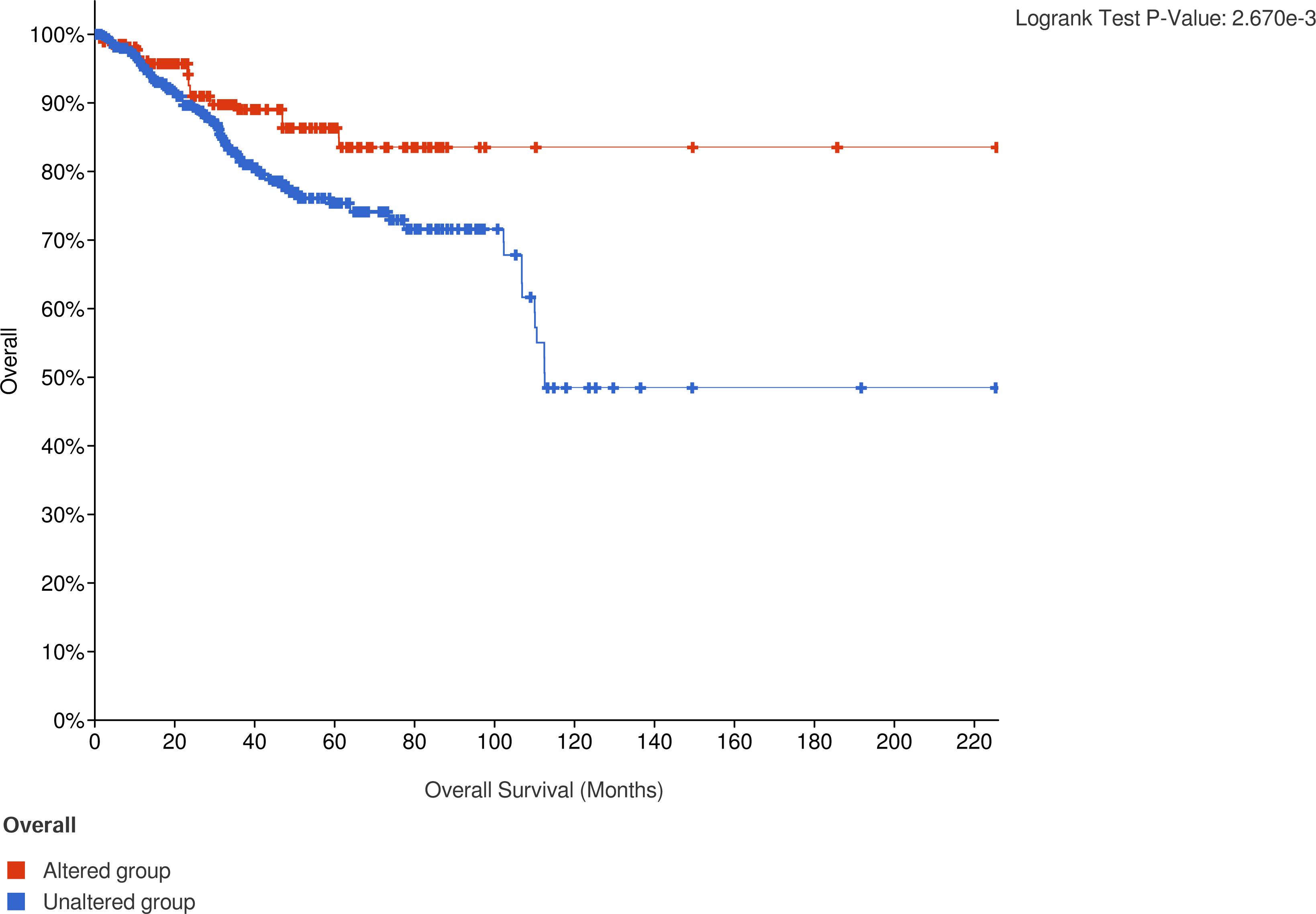
Overall survival by *CTNNB1* mutation status in the TCGA dataset (p<0.05).

In a multivariate analysis for OS and RFS including MSI, POLE-Status, tumor grade and tumor stage, this effect was lost for β-catenin as well as for LEF1 - only tumor grade and stage remained significant.

In addition to *CTNNB1*, eighteen other tumors showed mutations of the WNT signaling pathway (Supplementary table 1). LEF1 correlated significantly with all mutations of the wnt pathway tested (p<0.001) as did β-catenin (p<0.001).

### β-catenin and LEF1 in diagnostic setting of fibromatoses and other tumor types with recurrent *CTNNB1* mutations

Fibromatoses of the desmoid-type carried *CTNNB1* mutations in 63% of cases (5/8). β-catenin and LEF1 were similarly positively expressed in all mutant and two wild-type cases. Superficial fibromatoses were all *CTNNB1* wild-type and immunohistochemically, β-catenin and LEF1 were again similarly expressed (positive in one, negative in two cases). The LEF1 readout was easier in all cases (Fig. 5 a-c). In addition, we applied LEF1 immunostaining in addition β-catenin in a variety of other tumor types that we encountered in our routine practice (including solid pseudopapillary tumors of the pancreas, a pancreatoblastoma, pilomatricomas, medulloblastomas and adamantinomatous craniopharyngiomas) known to harbor a *CTNNB1* mutation respectively (data not shown). Again, we found LEF1 to be consistently easier to evaluate, usually the result being evident at low magnification (Fig. 5 d-f).

**Figure 5:**
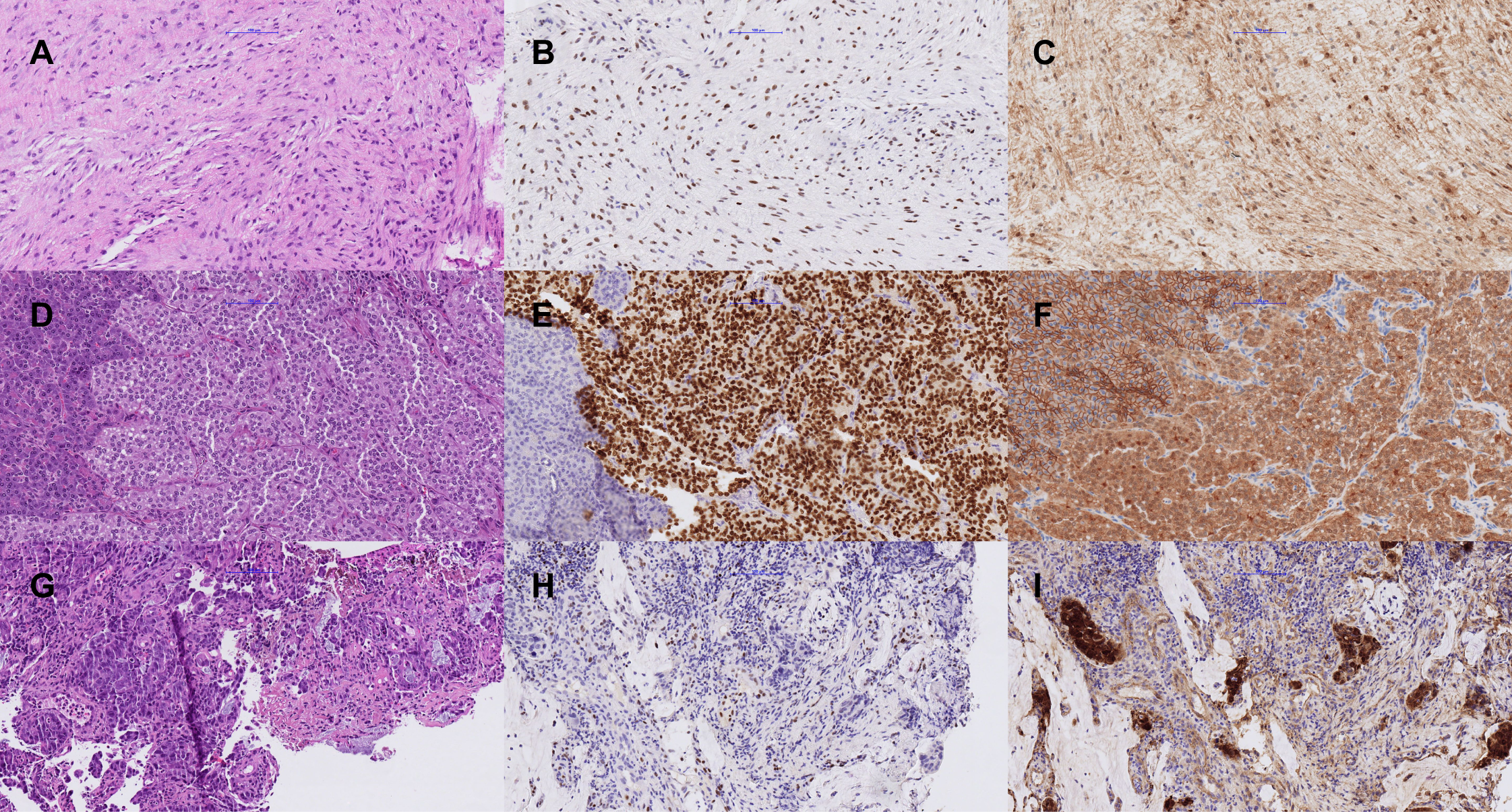
A-C: Desmoid type fibromatosis: A: HE showing classic bland spindle cell morphology; B: nuclear positivity of LEF1, C: nuclear and cytoplasmic positivity of β-catenin. D-F: Solid pseudopapillary tumor of the pancreas (SPN): dD: HE staining, SPN on the right side, normal pancreas on the left; E: nuclear positivity of LEF1 in SPN while normal pancreas is negative, F: nuclear and cytoplasmic positivity of β-catenin in SPN as compared to membranous positivity in normal pancreatic tissue. G-I: Intestinal adenocarcinoma: G: intestinal differentiated adenocarcinoma; H: infiltrative carcinoma is negative for LEF1; I: strong nuclear and cytoplasmic positivity of the adenocarcinoma for β-catenin.

### β-catenin and LEF1 in various neoplasms

*CTNNB1* mutations were incidentally detected in the context of NGS performed within diagnostic routine across a variety of tumor types (adenocarcinoma of the lung n=5, colon carcinoma n=5, malignant melanoma n=3, adrenocortical carcinoma, prostate carcinoma, pancreatic adenocarcinoma, medulloblastoma, neuroendocrine carcinoma of the lung) in a total of 18 cases (Supplementary Table 2). Immunohistochemistry for β-catenin predicted mutational status correctly in 89% of cases, whereas LEF1 only in 50%. When another strong driver mutation such as *KRAS, BRAF, EGFR* or *TP53* was present at the same time, the mutation status of β-catenin was correctly predicted in 87%, while LEF1 was correctly predicted in 53% (Supplementary Table 2).

## Discussion

We investigated the utility of LEF1 immunohistochemistry as a surrogate marker for *CTNNB1* mutations in various human neoplasms in different diagnostic and prognostic settings. Overexpression of LEF1 has been described in several tumor types harboring *CTNNB1* mutations, such as cribriform-morular thyroid carcinoma [15,16] or deep penetrating melanocytic naevi [17], but has not been systematically studied as surrogate for *CTNNB1* mutations across different tumor types.

We found that LEF1 is useful as a surrogate of *CTNNB1* mutations across various different entities. In a large cohort of endometrial carcinomas, we found LEF1 to be slightly less sensitive than β-catenin (64% vs 72%) but markedly more specific (95% vs 76%). Perhaps more importantly, we found that LEF1 was easier to interpret than β-catenin and that most cases were easy to assess at low magnification. This is due to the simple readout of LEF1 immunostaining (diffuse nuclear overexpression vs. its absence), while it is often difficult to distinguish between spillover of membranous β-catenin staining and true nuclear translocation, in particular as the latter may be only very focal. Furthermore, LEF1 benefits from the presence of internal positive controls (in particular T cells) in essentially all tissues of interest. We furthermore corroborated these findings across a number of tumor types.

A possible limitation of the immunohistochemical results gained in the cohort of endometrial carcinomas relates to the use of tissue microarray with regard to intratumoral heterogeneity. However, our preliminary results obtained with full histological slides and the use of three punches per case, as well as the correlation with the molecular results, suggest that the findings would not have been substantially different with staining on whole tissue sections.

### β-catenin and LEF1 as a diagnostic marker

*CTNNB1* mutations can be found in the majority of sporadic fibromatoses [18]. They are known to carry this mutation in about 90% of cases which is used diagnostically with β-catenin immunohistochemistry as a surrogate marker. Normally, β-catenin staining patterns are membranous and cytoplasmic. In case of a *CTNNB1* mutation or canonical WNT pathway activation, β-catenin is translocated to the nucleus where it activates downstream transcriptional programs [19]. As previously reported, we could identify a nuclear positivity of β-catenin in most deep fibromatoses. What is new is that we also stained for LEF1, which is downstream of β-catenin, and here we saw that this marker was far easier to evaluate in all cases. Interestingly, this holds also true for superficial fibromatoses which do not carry a *CTNNB1* mutation but nevertheless show nuclear positivity of β-catenin [20,21]. A recent study assessed the accuracy of three different β-catenin clones for prediction of *CTNNB1* mutation status in suspected desmoid-type fibromatosis and found a variation regarding both sensitivity and specificity [22]. AThe antibody clone 14 used in the present study showed the highest sensitivity (96%) and the lowest specificity (62%) in this latter study. The authors furthermore assessed LEF1 immunostaining using the same clone used in the present study and found it to be 88% sensitive and 76% specific. recent study applying the LEF1 clone EP310 in desmoid-type fibromatosis found similar sensitivities and specificities for LEF1 and β-catenin alone, while the combination of both improved specificity [23].

Of note, neither nuclear β-catenin staining nor LEF1 overexpression were present in occasional cases with *CTNNB1* in addition to another strong driver. This finding is in line with published data for β-catenin immunostaining [5] and may suggest that the WNT pathway is not actually active in the presence of another potent tumor driver.

### Prognostic role of β-catenin and LEF1 in endometrial carcinomas

The role of β-catenin immunohistochemistry has been studied in the past. It was demonstrated that approximately 40% of endometrial carcinomas have nuclear β-catenin expression in less than 10% of tumor cells [5]. Although the authors concluded that β-catenin can reliably predict mutational status of *CTNNB1*, a marker that is somewhat easier to assess and does not require much expertise to obtain a reliable result would obviously be very helpful, which we found to be true for LEF1.

LEF1 was upregulated at the tumor front and in areas of presumable epithelial-mesenchymal transition which fits well in the concept of an activated WNT signaling pathway under these circumstances [24]. The pattern of immunostaining differed however from *CTNNB1* mutation-associated LEF1 overexpression in that it was focal, variably intense and would not be present in the majority of tumor cells.

Ruz-Caracuel et al. compared the prediction of *CTNNB1* exon 3 mutations by immunohistochemistry for β-catenin and LEF-1 in low-grade, early-stage endometrial endometrioid carcinoma and found β-catenin had a higher predictive value than LEF1 for these mutations [25]. This difference to our findings might be due to the use of different clones for both LEF1 (EP310) and β-catenin as well as a very low cutoff for LEF1 to be considered overexpressed (Allred score of 3/15). Indeed, the microphotographs provided in this article might suggest technical issues with LEF1 immunostaining compared to both our findings and to other studies using the same LEF1 clone EP310 [26,27]. An adverse effect of the presence of a *CTNNB1* mutation in these tumors has been reported as well [5,25]. A subgroup analysis of early endometrioid carcinomas in our cohort revealed a nonsignificant trend toward an unfavorable outcome in the presence of a *CTNNB1* mutation, confirming these results. However, when we included all carcinomas into the analysis, the presence of a *CTNNB1* mutation in our data was a favorable sign, which we also saw in the TCGA dataset [28].

Nevertheless, the difficulty of β-catenin immunohistochemistry persists and while one group does not comment on this topic, the other study provides an indirect hint as it reported only poor correlation between β-catenin and *CTNNB1* mutation [5], underscoring the need of a better surrogate marker.

The reliable identification of *CTNNB1* mutations by immunohistochemistry is also underscored by the recently published PORTEC-4a trial, in which endometrial carcinomas with mismatch repair deficits were classified into different risk groups based on their mutation status [4].

### Conclusion

We report immunohistochemical and prognostic results of LEF1 in a series of 130 endometrial carcinomas and compare them with β-catenin staining results. The *CTNNB1* mutation status serves as the ground truth in all cases. In addition, we are investigating the role of LEF1 in other diagnostically difficult lesions known to harbor CTNNB1 mutations, such as fibromatoses or a solid pseudopapillary tumor of the pancreas. Finally, we are investigating the role of LEF1 and β-catenin in a number of different malignant neoplasms, most of which have a different driver mutation concomitantly.

We show that LEF1 immunohistochemistry can be used as a diagnostic and predictive tool and can be used to predict *CTNNB1* mutations as well as β-catenin. And while the β-catenin readout requires a lot of expertise, the readout of LEF1 is much easier. The negativity of immunohistochemistry of β-catenin and LEF1 in the presence of a *CTNNB1* mutation and a strong known other driver mutation might indicate that the EMT pathway is not active, which may play a role in tumor progression and patient management.

## Data Availability

All data produced in the present study are available upon reasonable request to the authors

## Acknowledgements

The authors thank the staff of the Translational Research Unit at the Institute of Pathology, University of Bern, for excellent technical Support.

## Author contributions

MSD and EH conceived the study and wrote the manuscript. All authors analyzed data and approved the final version of the manuscript.

## References

1 Kubota Y, Kawakami H, Natsuizaka M, et al. CTNNB1 mutational analysis of solid-pseudopapillary neoplasms of the pancreas using endoscopic ultrasound-guided fine-needle aspiration and next-generation deep sequencing. J Gastroenterol. 2015;50:203–10.

2 Colombo C, Bolshakov S, Hajibashi S, et al. ‘Difficult to diagnose’ desmoid tumours: a potential role for CTNNB1 mutational analysis. Histopathology. 2011;59:336–40.

3 Carlson JW, Fletcher CDM. Immunohistochemistry for beta-catenin in the differential diagnosis of spindle cell lesions: analysis of a series and review of the literature. Histopathology. 2007;51:509–14.

4 Kurnit KC, Kim GN, Fellman BM, et al. CTNNB1 (beta-catenin) mutation identifies low grade, early stage endometrial cancer patients at increased risk of recurrence. Mod Pathol. 2017;30:1032–41.

5 Kim G, Kurnit KC, Djordjevic B, et al. Nuclear β-catenin localization and mutation of the CTNNB1 gene: a context-dependent association. Mod Pathol. 2018;31:1553–9.

6 Travaglino A, Raffone A, Saccone G, et al. Immunohistochemical Nuclear Expression of β-Catenin as a Surrogate of CTNNB1 Exon 3 Mutation in Endometrial Cancer. Am J Clin Pathol. 2019;151:529–38.

7 Constantine GD, Kessler G, Graham S, et al. Increased Incidence of Endometrial Cancer Following the Women’s Health Initiative: An Assessment of Risk Factors. J Womens Health. 2019;28:237–43.

8 van Aalten SM, Verheij J, Terkivatan T, et al. Validation of a liver adenoma classification system in a tertiary referral centre: implications for clinical practice. J Hepatol. 2011;55:120–5.

9 Menter T, Dirnhofer S, Tzankov A. LEF1: a highly specific marker for the diagnosis of chronic lymphocytic B cell leukaemia/small lymphocytic B cell lymphoma. J Clin Pathol. 2015;68:473–8.

10 Boos LA, Dettmer M, Schmitt A, et al. Diagnostic and prognostic implications of the PAX8-PPARγ translocation in thyroid carcinomas-a TMA-based study of 226 cases. Histopathology. 2013;63:234–41.

11 Rau TT, Bettschen E, Büchi C, et al. Prognostic impact of tumor budding in endometrial carcinoma within distinct molecular subgroups. Mod Pathol. 2021;34:222–32.

12 Hewer E, Phour J, Gutt-Will M, et al. TERT Promoter Mutation Analysis to Distinguish Glioma From Gliosis. J Neuropathol Exp Neurol. 2020;79:430–6.

13 Cerami E, Gao J, Dogrusoz U, et al. The cBio cancer genomics portal: an open platform for exploring multidimensional cancer genomics data. Cancer Discov. 2012;2:401–4.

14 Gao J, Aksoy BA, Dogrusoz U, et al. Integrative analysis of complex cancer genomics and clinical profiles using the cBioPortal. Sci Signal. 2013;6:l1.

15 Mohindra S, Sakr H, Sturgis C, et al. LEF-1 is a Sensitive Marker of Cribriform Morular Variant of Papillary Thyroid Carcinoma. Head Neck Pathol. 2018;12:455–62.

16 Dettmer MS, Hürlimann S, Scheuble L, et al. Cribriform Morular Thyroid Carcinoma - Ultimobranchial Pouch-Related? Deep Molecular Insights of a Unique Case. Endocr Pathol. Published Online First: 30 May 2023. doi: 10.1007/s12022-023-09775-z

17 Raghavan SS, Saleem A, Wang JY, et al. Diagnostic Utility of LEF1 Immunohistochemistry in Differentiating Deep Penetrating Nevi From Histologic Mimics. Am J Surg Pathol. 2020;44:1413–8.

18 Desmoid Tumor Working Group. The management of desmoid tumours: A joint global consensus-based guideline approach for adult and paediatric patients. Eur J Cancer. 2020;127:96–107.

19 Rubinfeld B, Robbins P, El-Gamil M, et al. Stabilization of beta-catenin by genetic defects in melanoma cell lines. Science. 1997;275:1790–2.

20 Montgomery E, Lee JH, Abraham SC, et al. Superficial fibromatoses are genetically distinct from deep fibromatoses. Mod Pathol. 2001;14:695–701.

21 Goto K, Ishikawa M, Aizawa D, et al. Nuclear β-catenin immunoexpression in scars. J Cutan Pathol. 2021;48:18–23.

22 Yamada Y, Hirata M, Sakamoto A, et al. A comparison of the usefulness of nuclear beta-catenin in the diagnosis of desmoid-type fibromatosis among commonly used anti-beta-catenin antibodies. Pathol Int. 2021;71:392–9.

23 Jobbagy S, Lozano-Calderon S, Mullen JT, et al. Utility of LEF1 to differentiate desmoid fibromatosis from its histologic mimics. Virchows Arch. 2024;484:807–13.

24 Bai Y, Sha J, Kanno T. The Role of Carcinogenesis-Related Biomarkers in the Wnt Pathway and Their Effects on Epithelial-Mesenchymal Transition (EMT) in Oral Squamous Cell Carcinoma. Cancers. 2020;12. doi: 10.3390/cancers12030555

25 Ruz-Caracuel I, López-Janeiro Á, Heredia-Soto V, et al. Clinicopathological features and prognostic significance of CTNNB1 mutation in low-grade, early-stage endometrial endometrioid carcinoma. Virchows Arch. Published Online First: 21 August 2021. doi: 10.1007/s00428-021-03176-5

26 Wang D, Gong J, Zhang H, et al. Immunohistochemical staining of LEF-1 is a useful marker for distinguishing WNT-activated medulloblastomas. Diagn Pathol. 2022;17:69.

27 Farzaneh T, Nowroozizadeh B, Han M, et al. [Diagnostic Utility of LEF1 Immunostain in Cytology Specimens of Solid Pseudopapillary Neoplasm of Pancreas]. Acta Cytol. 2021;65:250–6.

28 cBioPortal for Cancer Genomics. http://cbioportal.org (accessed 21 January 2022)

