## Supplementary Table 1 for "Lymphoid Enhancer-Binding Factor 1 (LEF1) immunostaining as a surrogate of β-catenin (*CTNNB1)* mutations"

| Gene | Alteration | Second Gene | Alteration |
| --- | --- | --- | --- |
| APC | p.R1096X p.S1447Wfs*3 |  |  |
| APC | p.S1178L |  |  |
| AXIN1 | p.G508Vfs*197 | AXIN2 | p.G589Afs*35 |
| AXIN2 | p.G600Afs*24 |  |  |
| AXIN2 | p.E698X |  |  |
| CSNK1A1 | p.R264C |  |  |
| CTNNB1 | p.D32N | APC | p.R2219X |
| CTNNB1 | p.G34V | EP300 | p.H2298Tfs*29 |
| CTNNB1 | p.S33C |  |  |
| CTNNB1 | p.S37Y |  |  |
| CTNNB1 | p.S33C |  |  |
| CTNNB1 | p.G34R |  |  |
| EP300 | p.R1266Gfs*11 |  |  |
| EP300 | p.Q498X |  |  |
| EP300 | splicesite |  |  |
| EP300 | p.R2291W |  |  |
| GSK3B | p.I238T |  |  |
| MYC | p.D135N |  |  |
| PPP2R1A | p.P179R |  |  |
| PPP2R1A | p.S77F |  |  |
| ppp2r1A | p.S40L |  |  |
| PPP2R1A | p.P179R |  |  |
| PPP2R1A | p.S77F |  |  |
| PPP2R1A | p.V80M |  |  |
| PPP2R1A | p.E101K |  |  |
| TCF7L2 | p.A544V |  |  |

Supplementary Table 1. WNT signaling pathway mutations other than *CTNNB1* mutations identified through panel sequencing.
