## Supplementary Table 2 for "Lymphoid Enhancer-Binding Factor 1 (LEF1) immunostaining as a surrogate of β-catenin (*CTNNB1)* mutations"

| **Tumour Type** | **Mutation CTNNB1** | **other Mutations** | **beta-Catenin IHC** | **LEF1 IHC** |
| --- | --- | --- | --- | --- |
| Adrenocortical carcinoma | CTNNB1 p.Ser37Pro (c.109T>C) | wt | positive | positive |
| Ampullary adenocarcinoma, intestinal type | CTNNB1 p.Thr41Ala (c.121A>G) | TP53 p.Met169Ter (c.505delA), TP53 p.Arg248Trp (c.742C>T) | positive | negative |
| Colon carcinoma | CTNNB1 p.Gly34Glu (c.101G>A) | BRAF p.Val600Glu (c.1799T>A), MAP2K1 p.Asp67Asn (c.199G>A), CDKN1B p.Lys25fs (c.72delC), FGF3 p.Thr136Met (c.407C>T), FGF3 p.Gly94Arg (c.280G>A), FANCA p.Val384Ile (c.1150G>A), SMAD4 p.Ala532Thr (c.1594G>A), SMARCA4 p.Ala1132Thr (c.3394G>A), NOTCH3 p.Glu1670Lys (c.5008G>A) | positive | negative |
| Colon carcinoma | CTNNB1 p.Thr41Ala (c.121A>G) | KRAS p.Gly12Asp (c.35G>A) | positive | negative |
| Colon carcinoma | CTNNB1 p.Gly34Glu (c.101G>A) | wt | positive | negative |
| Colon carcinoma | wt | APC p.Gln1406Ter (c.4216C>T) | positive | positive |
| Colon carcinoma | CTNNB1 p.Ser45Pro (c.133T>C) | PIK3CA p.His1047Arg (c.3140A>G), DDR2 p.Arg105His (c.314G>A) | positive | positive |
| Large cell neuroendocrine carcinoma of the lung | CTNNB1 p.Ser33Phe (c.98C>T) | CREBBP p.Ser944Ter (c.2831C>G), TP53 p.Gln192Ter (c.574C>T) | positive | positive |
| Lung Adenocarcinoma | CTNNB1 p.Gly34Glu (c.101G>A) | EGFR p.Glu746_Ala750del (c.2235_2249del) | negative | negative |
| Lung Adenocarcinoma | CTNNB1 p.Pro44Leu (c.131C>T) | KRAS p.G13_V14>DI (c.38_40GCG>ACA) | negative | negative |
| Lung Adenocarcinoma | CTNNB1 p.Ser33Phe (c.98C>T) | EGFR p.Glu746_Ala750del (c.2235_2249delGGAATTAAGAGAAGC), TP53 p.Glu298Ter (c.892G>T) | positive | negative |
| Lung Adenocarcinoma | CTNNB1 p.Asp32Asn (c.94G>A) | KRAS p.Gly12Asp (c.35G>A) | positive | negative |
| Lung Adenocarcinoma | CTNNB1 p.Ser37Phe (c.110C>T) | KRAS p.Gly12Val (c.35G>T) | positive | negative |
| Malignant melanoma | CTNNB1 p.Ser37Tyr (c.110C>A) | BRAF p.Val600Lys (c.1798_1799delGTinsAA), PIK3CA p.Gly320Arg (c.958G>A), FGFR3 p.Pro250Leu (c.748_749delCCinsTT) | positive | positive |
| Malignant melanoma | CTNNB1 p.Asp32Tyr (c.94G>T) | KIT p.Thr574_Tyr578delinsAsn (c.1721_1732delCACAACTTCCTT) | positive | positive |
| Malignant melanoma | CTNNB1 p.Asp32Asn (c.94G>A) | NRAS p.Gln61Lys (c.181C>A) | positive | positive |
| Medulloblastoma | CTNNB1 p.Asp32Tyr (c.94G>T) | ALK p.Ile1183Met (c.3549T>G), RB1 p.Ser187Phe (c.560C>T) | positive | positive |
| Prostate adenocarcinoma | CTNNB1 p.Ser45Pro (c.133T>C) | wt | positive | positive |

Supplementary Table 2. Sequencing and immunostaining findings for cases with incidentally detected WNT pathway mutations.
